# Giving a voice to adults with COVID-19: An analysis of open-ended comments from smell longhaulers and non-longhaulers

**DOI:** 10.1101/2023.03.07.23286910

**Authors:** N. S. Menger, A. Tognetti, M. C. Farruggia, C. Mucignat, S. Bhutani, K. W. Cooper, P. Rohlfs Dominguez, T. Heinbockel, V. Shields, A. D’Errico, V. Pereda-Loth, D. Pierron, S. Koyama, I. Croijmans

## Abstract

Smell disorders are commonly reported with COVID-19 infection. Some patients show prolonged smell-related issues, even after the respiratory symptoms are resolved. To explore the concerns of patients, and to provide an overview for each specific smell disorder, we explored the longitudinal survey that was conducted by ^1^, and contained self-reports on the changes of smell that participants experienced at two time points. People who still suffered from smell disorders at the second time point, hence named ‘longhaulers’, were compared to those who were not, hence named ‘non-longhaulers’. Specifically, three aims were pursued in this study. First, to classify smell disorders based on the participants’ self-reports. Second, to classify the sentiment of each self-report using a machine learning approach, and third, to find specific keywords that best describe the smell dysfunction in those self-reports. We found that the prevalence of parosmia and hyposmia was higher in longhaulers than in non-longhaulers. Furthermore, the results suggest that longhaulers stated self-reports with more negative sentiment than non-longhaulers. Finally, we found specific keywords that were more typical for either longhaulers compared to non-longhaulers. Taken together, our work shows consistent findings with previous studies, while at the same time, provides new insights for future studies investigating smell disorders.

## Introduction

Chemosensory dysfunctions are among the distinguishing symptoms of COVID-19^2,3^. While many infected patients recover within weeks, a large percentage experiences long-term olfactory dysfunction even after recovery from the acute phase^4–6^. These individuals are known as *smell longhaulers*. Since smell impairment is often hard to notice, and even harder to describe by patients (e.g. patients commonly confuse smell disorders with taste disorders), information that can clarify the process of smell impairment or recovery can be useful to describe, understand, and track the phenomenon (see ^7^). It is also unclear how distorted chemosensory perception relates to well-being or changes in behavior, such as those related to food intake or avoidance. The present study aims to address this research gap by applying a set of methods taken from natural language processing and linguistics to open-ended responses collected in a large-scale survey^1^. Our analysis provides a new perspective on olfactory disturbances following COVID-19 that cannot be captured through closed-ended questions. On a more general level, this data science perspective can advance online survey-based patient research studies.

Disturbances of smell can be classified as either quantitative or qualitative. Quantitative changes can occur when the intensity of perceived odors is altered; these can range from an increased perceived intensity (hyperosmia) to a more faint perception of the odor (hyposmia) or even a complete loss of olfactory perception (anosmia). On the other hand, qualitative changes may occur when the quality of the perceived odor is altered^8,9^, which occurs, for instance, in case of parosmia (i.e., distorted smell perception) or phantosmia (i.e., smelling things that are not present). Qualitative smell changes, such as parosmia and phantosmia are often reported in smell longhaulers after COVID-19 and in people who report recovered or improved quantitative smell changes^1,10^ (i.e., anosmia or hyposmia). While these smell distortions are common, most people are unaware of the technical terminology for such distortions, but may nevertheless be able to describe their condition in an open-ended format. Confusion may for example arise from terms like “flavor” and “taste”^11^, where individuals may report that they do not taste anything while only their smell is affected and not their taste. Similarly, different types of smell changes may cause confusion, as the quantity (complete or partial loss of smell) and quality of the changes indicate different impairments. Moreover, people are often inaccurate in reporting their olfactory performances, which may affect their emotional well-being and their awareness of olfactory dysfunction^12^. It is important to correctly classify symptoms of smell disorders, both for clinical tracing and potential treatment choice, as well as for scientific research purposes^13^ (i.e., to better define each type of chemosensory dysfunction). These difficulties in symptom description make it hard to capture a person’s specific olfactory experience in close-ended survey questions^10,14,15^ versus open-ended questions. In this study, we used an open-ended survey, which offered respondents the opportunity to provide comments^2^.

There is a strong link between the sense of smell and a person’s emotional and cognitive states related to the connections between olfactory and emotional processing areas of the brain. The cortical nucleus of the amygdala, an area involved in olfactory processing, receives direct input from the olfactory bulb^16^. The amygdala is linked to the control and regulation of emotions and social cognition^17^. The mediodorsal nucleus of the thalamus (MDT) receives olfactory input from the olfactory cortex, from brain areas that receive direct monosynaptic input from the olfactory bulb (OB). These, in turn, project to the orbitofrontal cortex (OFC), a region that is important for executive functions and integrating olfactory and gustatory inputs (and thus flavor perception), compiling information regarding reward value, decision making^16^, and in memory. The primary olfactory cortex also sends projections to the hippocampus^18^, which is mainly involved in memory processes through the rostral entorhinal cortex^19^. Connections between the entorhinal cortex and the hippocampus and neocortical regions are reciprocal^19^. With each sniff, all of these neural connections are activated^20^. Moreover, olfactory dysfunctions are present in, and often precede, neurodegenerative disorders, such as Alzheimer’s and Parkinson’s disease^21^, autism, where social cognition is affected^17^, and in depression, where mood is affected^22^. At the same time, emotional and cognitive deficits are shown by patients with olfactory dysfunction even several months after experiencing mild, moderate, or severe COVID-19 disease^20,23^. Therefore, emotional and cognitive well-being might be affected by olfactory dysfunction. The detrimental effect of chemosensory dysfunction on emotional well-being is well recognized^24,25^ (see ^26^ for a review), but is not fully understood with respect to olfaction in longhaulers. Patients have reported altered mental status, as well as frustrations with COVID-19-related olfactory dysfunction^27^. Furthermore, olfactory dysfunction predicts the development of depression in adults^22^. Therefore, a negatively affected well-being or emotional tone when describing their symptoms would be expected in patients with COVID-19 and olfactory dysfunctions, such as anosmia or parosmia, particularly in longhaulers.

In the present work, we analyzed open-ended questions that were included in surveys conducted by the Global Consortium for Chemosensory Research (GCCR)^2,3^. As an initial validation of the informative value of the comments, a comparison was made between symptoms coded from open ended comments and from the multiple choice answers alone, administered during COVID-19 (S1) and after recovery (S2). This set the stage to address the following three questions: (1) What are the frequencies of parosmia, phantosmia, and other olfactory dysfunctions (i.e. hyposmia and anosmia) as reported in open-ended comments? (2) What is the well-being or emotional tone of people suffering from these symptoms as reported in open-ended comments? (3) What specific food-related experiences are related to these symptoms? Open-ended questions allow participants to voice their concerns that may not be covered by the other type of questions, and are closer to how patients may report these symptoms to their general practitioner or health-care worker^28^. The questions addressed smell loss while participants experienced COVID-19 (S1) and during a follow-up survey (S2). Analysing these comments and their content contributes to a better understanding, in a more ecologically valid way, of how longhauling might affect emotional well-being as it relates to olfactory experiences and the frequency and severity of symptoms compared with analysing close-ended survey questions alone. On the basis of previously reported information, the following hypotheses have been formulated in the present study.

### Hypothesis Aim 1

Recovery from smell loss is often accompanied by parosmia and phantosmia, and is considered a sign of olfactory mucosa regeneration (e.g. ^1^). Considering that some smell-related symptoms may remain in COVID-19 longhaulers, we predict that longhaulers will have a greater occurrence of parosmia and phantosmia in addition to other potential chemosensory dysfunctions, compared with non-longhaulers based on their own description of their olfactory symptom progression at S2.

### Hypothesis Aim 2

Using a machine learning aspect-based sentiment analysis, we predict that longhaulers will report significantly more emotional and psychological distress compared to non-longhaulers.

### Hypothesis Aim 3

We hypothesize that longhaulers reporting parosmia and phantosmia will exhibit avoidance behaviour, resulting in omission of certain food and non-food items. This will be apparent from a qualitative semantic analysis of the comments at S2.

## Methods

This work has been designed and planned following the structure of a survey and used data previously available to the GCCR^2,3^ that analyzed the closed-ended responses. More specifically, we used data acquired by means of open-ended questions included in those surveys. The existing protocol complies with the revised Declaration of Helsinki and was approved as an exempt study by the Office of Research Protections at The Pennsylvania State University (PennState) in the United States (STUDY00014904).

### Participants

The sample size of participants used in this study was 1560. We included respondents that completed the comments section in English, Dutch, French, Italian, or Spanish, from a second GCCR survey^3^, which was a continuation of an initial GCCR study^2^. This second survey was sent out to all participants that completed the first GCCR survey. Participants completed the second survey between 23 and 291 (median: 200) days after the first. The participants were classified as either non-longhauler or longhauler, based on their self-reported smell ability at S2 relative to S1. We refer the reader to ^1^ for a detailed overview of the data collection. Dutch, French, Italian, and Spanish comments were translated to English for our analyses. Translations were conducted by native speakers.

### Procedure

#### Aim 1

In previous text analyses^1,10^, an observation was made that longhaulers, but also some apparently recovered non-longhaulers, report sensations of (imagined) burning/unpleasant smells or altered smell and taste experiences. Our first aim was to determine the incidence of anosmia, hyposmia, phantosmia, and parosmia in longhaulers versus non-longhaulers by analyzing the free-text comments. At variance to ^1^, these complaints were categorized and counted based on descriptive comments and not on the participants’ self-reports by means of closed questions. The self-report question in ^1^ asking for changes in smell was meant to capture quantitative changes, and was not always sensitive to capture individual experiences. The question that prompted the free-text comment was “Please describe any current changes in smell. Type ‘none’ if this is not applicable”.

The data was first processed by means of a concept-driven quantitative content analysis, as described by^29^. Comments recorded at both time points (n=2543) were coded manually by eight different coders for the presence of anosmia, hyposmia, parosmia, phantosmia, whether the person indicated that they had recovered, not at all, partially or fully, and for mentioning food and non-food odorous items according to a predetermined coding scheme (see Coding scheme S1 in the Supplementary material, SM). Briefly, coders were first trained on the coding scheme by developing a set of rules, and a small subset of comments were coded followed by a discussion of comments that were difficult to code. Data was divided into eight roughly equal groups of around 350 comments. Groups were formed based on a semi-random number generator (draw 2542 times a number 1-8). A small set of comments (n=40), was coded by every coder to allow calculation of inter-coder agreement. Agreement between coders was calculated by means of Fleiss’s kappa^30^. Agreement between the eight coders was deemed acceptable, for anosmia (κ = .766), parosmia (κ = .777), phantosmia (κ = .575), hyposmia (κ = .691), presence of food items (κ = .796), and non-food items (κ = .740). No agreement was reached on whether people recovered fully, partially or not at all (κ = .127). Coders marked cases of which they were uncertain, and these were discussed during a group meeting. This discussion led to the inclusion of an additional variable, “hyperosmia”, or an *increased* sensitivity in the sense of smell. However, due to its low prevalence (n=13) none of the analyses included this variable. There were 169 cases that were unresolved, and these were again re-coded by two previous coders and two new coders. Unresolved comments from the two previous coders were exclusively analysed by the two new coders. Cases that did not meet any of the symptoms were excluded from further analyses.

Next, as an additional validation measure, the coded symptom prevalence of the free-text comments was compared to the incidence reported on the multiple-choice question that was used in ^1^. Here, participants were asked to rate whether they had noticed anosmia, hyposmia, parosmia or phantosmia by asking the following questions, respectively “*I cannot smell at all/ Smells smell less strong than they did before my impairment*” (anosmia/hyposmia), “*Smells smell different than they did before my impairment (the quality of smell has changed)*” (parosmia), and “*I can smell things that aren’t there (for example I smell burning when nothing is on fire)*” (phantosmia). Since multiple-choice questions for anosmia and hyposmia were not provided separately, we could only validate parosmia, phantosmia, as well as anosmia and hyposmia together.

#### Aim 2

Smell and taste disorders can lead to emotional distress (e.g. ^1,14^). Our first sub-aim was to determine if longhaulers demonstrated a greater reporting of emotional and psychological distress compared to non-longhaulers. Our second sub-aim was to examine if the emotional distress was linked with specific olfactory dysfunctions (e.g., phantosmia, parosmia). We therefore trained an aspect-based sentiment classification algorithm (LCF-ATEPC) from PyABSA^31^ (version 1.16.15) on a dataset of restaurant reviews available in PyABSA (“Restaurant16”) and then used the trained algorithm to first extract so-called aspects, i.e., keywords in each comment. The model examines what sentiment (negative, positive or neutral) is attributed to these aspects. Thus, more than one sentiment classification can be contained in one comment. Since the comments sometimes consisted of multiple sentences, this approach seemed more appropriate than aggregating the sentiment of each comment. To validate the results of the first model, we trained a second model with the same parameters on a dataset of laptop reviews also available in PyABSA (“Laptop14”), and used that model to classify the comments, and conduct the same analyses.

#### Aim 3

Once the incidence and impact of smell disorders was established, we determined whether specific foods, drinks or other objects were associated with longhauling, with the goal to examine whether specific items were more salient in longhaulers than non-longhaulers. We first extracted all food and non-food items from the comments that had been coded for aim 1. All items were visualized using word clouds. We then conducted a relative frequency analysis on the extracted food and non-food words from comments.

### Statistical analyses

#### Aim 1

For the validation of our coding, we computed the overlap between our coding of comments and the participants’ self-report. For each olfactory dysfunction, a confusion matrix was calculated. F-scores were then calculated for each matrix to indicate the extent of overlap.

We then used logistic regressions to approach Aim 1 (*glm* function with a binomial error structure of the *stats* package in R) and assessed whether the two categories of participants (long- vs. non-longhaulers) differed in terms of reported disorders they respectively experienced. The dependent variables were each of the four smell disorders studied, namely parosmia, phantosmia, hyposmia or anosmia (0 for absence, 1 otherwise). Our explanatory variable was “smell longhaulers status” (long- *vs* non-longhaulers). We included ‘age’ and ‘gender’ of the participants and whether the comments were translated in English or not (0 for untranslated comments *vs* 1 translated) as control variables as well as their interactions with the explanatory variable. We included all the main effects and interaction terms in the initial model, which was then simplified by removing the non-significant interaction terms to achieve the minimal adequate model. We centred ‘age’ in all the models in order to make the effects more easily interpretable. Statistical analyses were performed using R, version 4.1.3.

#### Aim 2

A chi-square test was conducted to examine the relationship between sentiment classifications and longhauler status of the aspect-based sentiment classification algorithm. A mixed-effects logistic regression model was used to examine the relationship of sentiment, longhauler status, and olfactory dysfunction type. All analyses were conducted with Python, version 3.8.8, and the packages Scipy, version 1.9.3, and Statsmodels, version 0.13.5.

#### Aim 3

This aim was approached by creating word clouds, and conducting a relative frequency analysis. For the word clouds, the extracted words describing food and non-food objects were converted to lower case unigrams, bigrams, and trigrams, using the R package RWeka (version 0.4.44) and TM (version 0.7.8). Using the R package wordcloud2 (version 0.2.1) and RColorBrewer (version 1.1.3), word clouds were created where the frequency of an n-gram determines the size within the ‘cloud’.

For the relative frequency analysis, we preprocessed and aggregated each group’s comments into a corpus based on longhauler status. Preprocessing steps included splitting the plain text comments into tokens. Tokens were lowercased, and numerals and punctuation removed. Commonly used stopwords were removed, and the text was lemmatized using Wordnet^32^. We then computed frequency lists for each corpus in the comparison based on our pre-processed comments. The log likelihood statistic was calculated for each word in the two frequency lists by constructing a contingency table based on word frequencies within and across corpora as per the method in ^33^. Given that log-likelihood is a statistical significance measure, it does not compute the size of the difference between corpora, rather, it provides the words we have most evidence for. Thus, to determine the influence of each word in each of the corpora, the relative frequency^34^ method was used. By comparing the normalised frequencies for each word, this method returns a value, [-1, 1]. In our case, 1 indicates that the word is overused in the longhauler corpus, and -1 in the non-longhauler corpus. Using these metrics, food and non-food words in the corpora were determined by manually coding words from each category with log likelihood values greater than 3.84 (a significance threshold of *p* < .05 or lower), and selected for further analysis.

## Results

The following section provides a description of the results for each aim. We also need to briefly address an important property of the comments, namely, many of them only contained the word “None”. The underlying reason to include “None,” was to give participants a voice who found that there were no changes in their smell. The responses that contained “None” entries (n=469) were excluded from further analyses.

As an additional validation measure, we examined the multiple choice question where participants had to report whether they experienced smell loss, parosmia or phantosmia. The F-score was calculated from confusion matrices (Table S2-S4 in the Supplementary material) for each smell disorder and served as an indicator of the agreement between coder and participant. The F-score for parosmia was 0.502, for phantosmia 0.416. The variable smell loss in the multiple choice question was compared to the coding of anosmia and hyposmia combined in the open-ended comments and had an F-score of 0.717.

### Aim 1

The logistic regression examining the association between anosmia and longhauler status could not be performed as the number of participants who reported to be suffering from anosmia was too small in both long- (17 out of 750 individuals) and non-longhaulers (0 out of 338 individuals; see Table 1 and Figure 1). For the other smell disorders, the minimal adequate models that were ran to obtain the results reported below, were obtained by removing the non-significant interaction terms between smell longhauler status and the three control variables (age, gender and translation), in all models (.062 < *p* < .983). The logistic regressions revealed a significant effect of the smell longhauler status (longhauler vs. non-longhaulers) in terms of reported disorders (Figure S8 in the SM): Longhauling participants were significantly more likely to report symptoms interpreted as parosmia (β = 0.58, SE = 0.14, *z* = 4.06, *p* < .001, 95% CI = 1.35-2.37, OR = 1.78, Table S5 in the SM) and hyposmia (β = 0.55, SE = 0.13, *z* = 4.16, *p* < .001, 95% CI = 1.34-2.26, OR = 1.74, Table S6 in the SM) compared to non-longhaulers. However, longhauler status did not affect the likelihood to report symptoms associated with phantosmia (β = 0.09, SE = 0.19, *z* = 0.49, *p* = .63, 95% CI = 0.76-1.62, OR = 1.10, Table S7 in the SM).

**Table 1.**
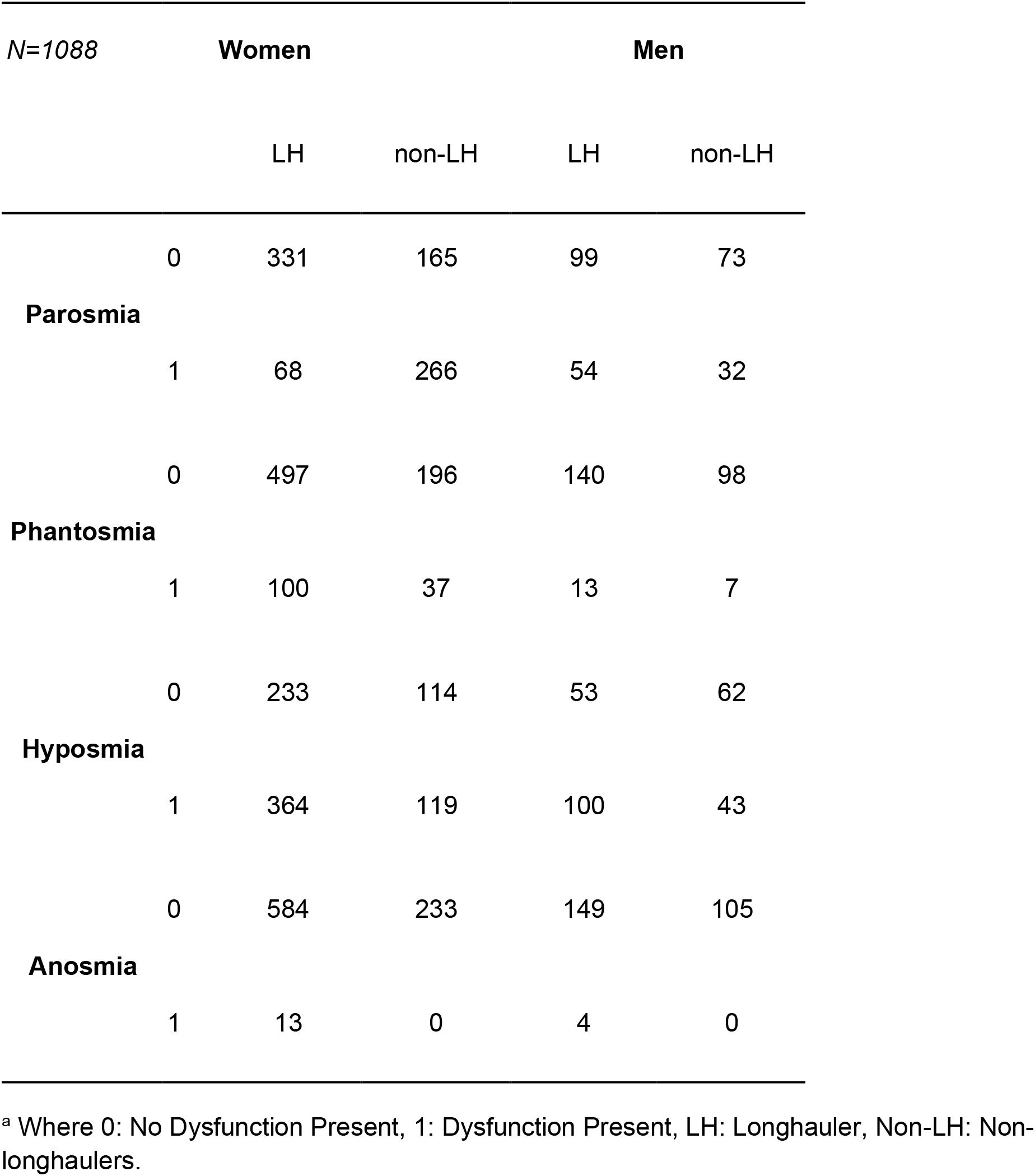
Sample Size of Reported olfactory dysfunctions by Smell Longhauling Status^a^

**Figure 1.**
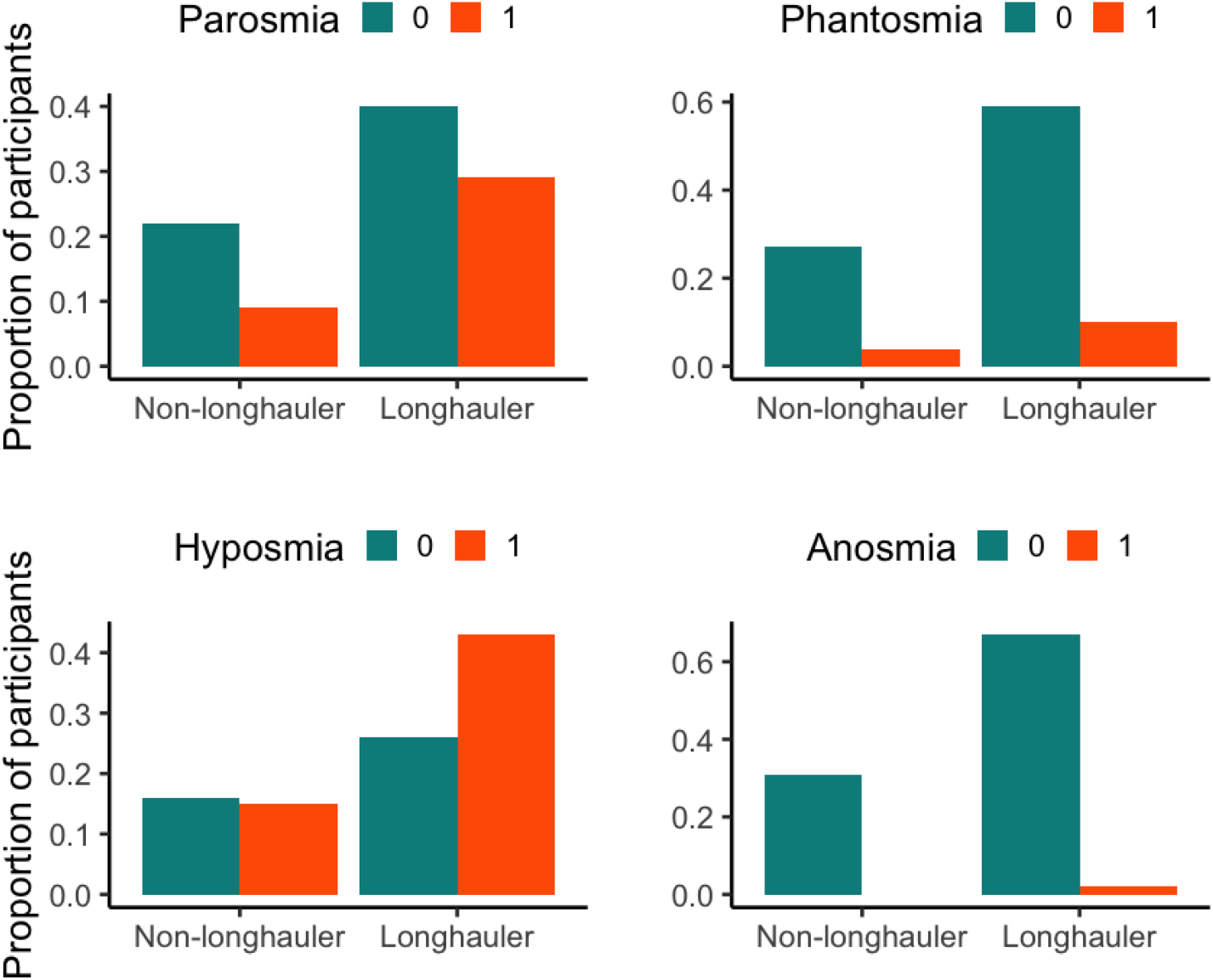
Prevalence of olfactory dysfunctions in longhaulers and non-longhaulers Proportion of participants with (orange) and without (turquoise) each olfactory dysfunction studied for both smell longhaulers and non-longhaulers.

### Aim 2

The aspects in each comment were classified as being negative, neutral or positive (numerically coded as 1, 2, 3, respectively). The classifier that was trained on restaurant reviews was unable to classify the sentiment of 854 out of 1560 comments, and 838 out of 1560 could not be classified by the model trained on laptop reviews. These comments were removed from further analyses. There was a relationship between the classified sentiment and longhauler status based on the model that was trained on restaurant reviews (*X*^2^ = 15.10, *p* < .001; Table 2) and laptop reviews (*X*^2^ = 30.32, *p* < .001).

**Table 2.**
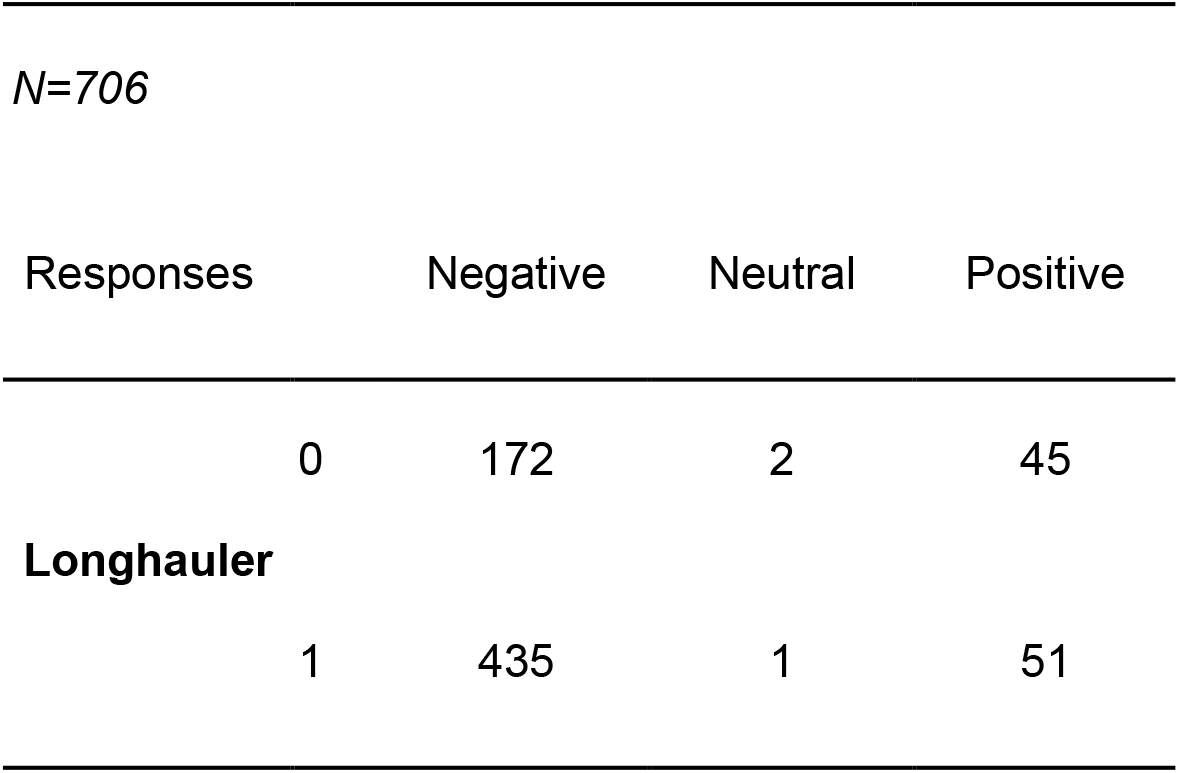
Sample Size of Classified Sentiment by Smell Longhauling Status for The Model Trained on Restaurant Reviews.

In addition to comparing the sentiment of longhaulers and non-longhaulers, an additional analysis was conducted to examine the effect of specific olfactory dysfunction on the comment’s sentiment. Within the comments of longhaulers (Figure 2) there was an effect of parosmia (β = -1.13, SE = 0.35, *p* = .001, 95% CI = 0.16-0.62, OR = 0.32), and hyposmia (β = -0.76, SE = 0.33, *p* = .023, 95% CI = 0.24-0.90, OR = 0.47). No significant effects were found for phantosmia (β = -0.53, SE = 0.50, *p* = .29, 95% CI = 0.19-1.45, OR = 0.59) and anosmia (β = -0.97, SE = 1.10, *p* = .38, 95% CI = 0.02-2.29, OR = 0.38).

**Figure 2.**
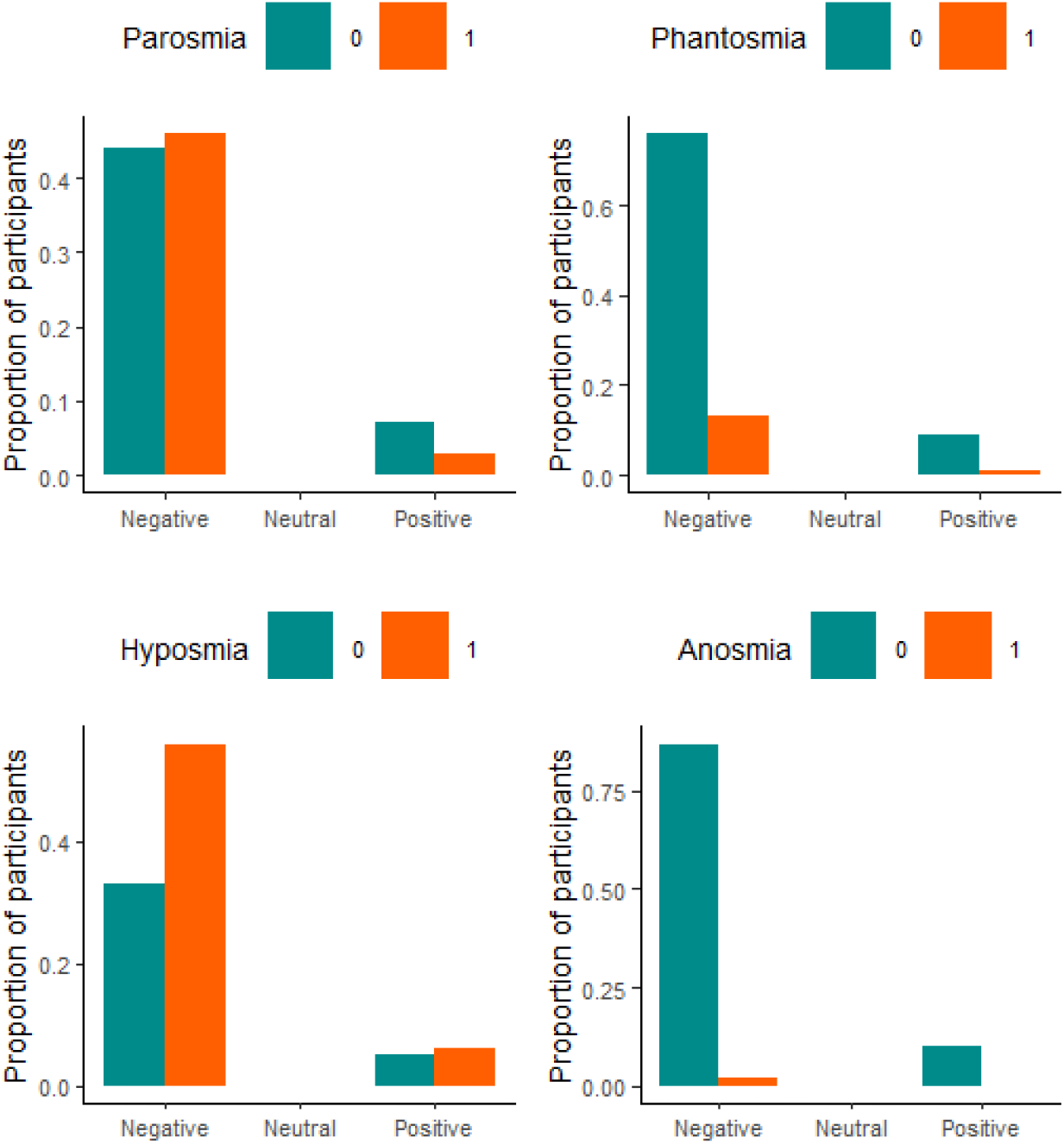
Sentiment for each olfactory dysfunction in longhaulers Proportion of longhaulers’ comments and their sentiment, as classified by the model trained on restaurant reviews.

For the analysis of non-longhaulers’ comments, the variable anosmia was omitted because none of the comments belonged to anosmic participants. In the non-longhauler comments, there was no significant difference between the classified sentiments (Figure 3). However, there was a significant effect of parosmia (β = -2.00, SE = 0.47, *p* < .001, 95% CI = 0.05-0.33, OR = 0.14) and hyposmia (β = -1.59, SE = 0.39, *p* < .001, 95% CI = 0.09-0.43, OR = 0.20), but not for phantosmia (β = -0.01, SE = 0.54, *p* = .98, 95% CI = 0.32-2.78, OR = 0.99).

**Figure 3.**
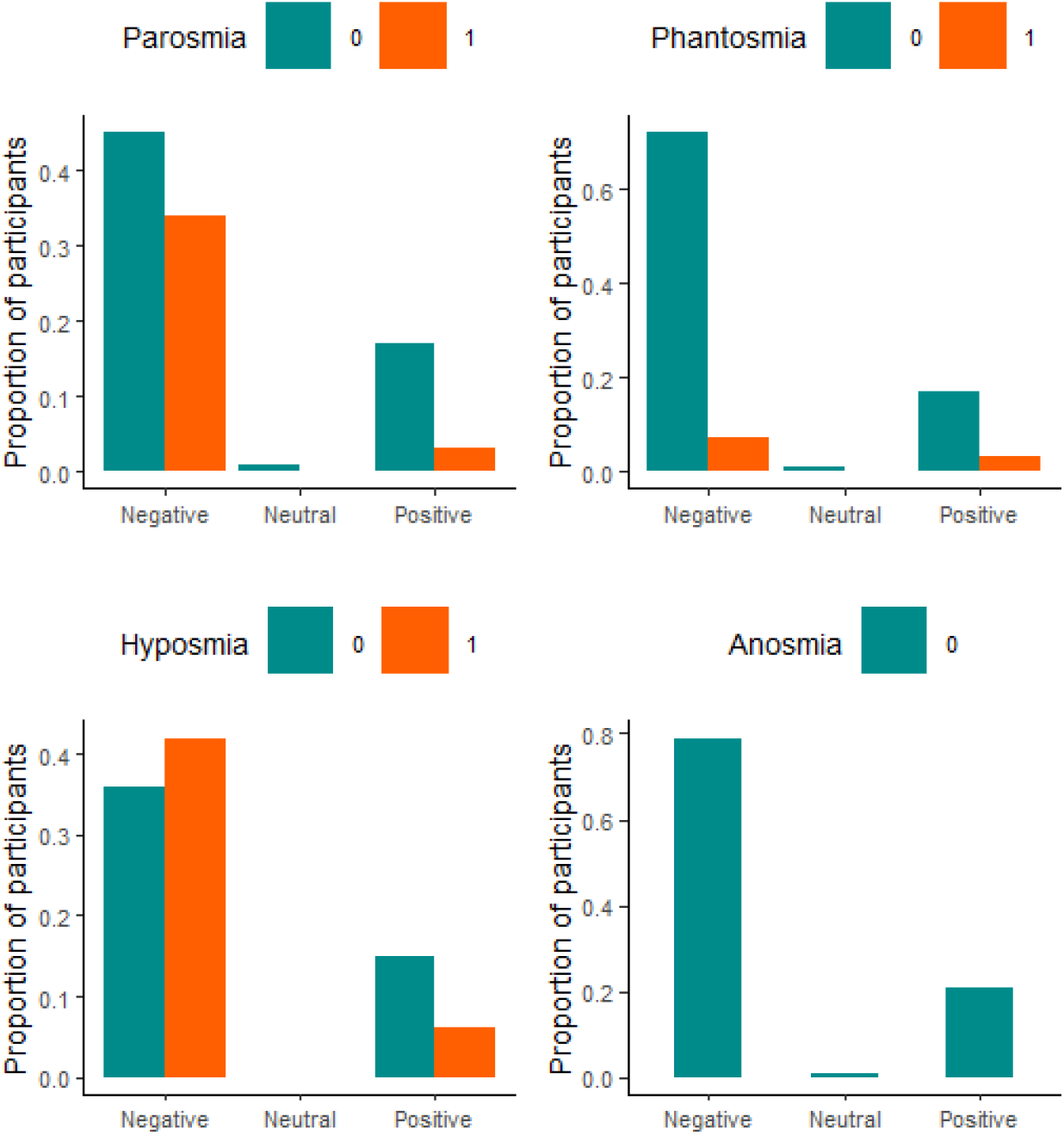
Sentiment for each olfactory dysfunction in non-longhaulers Proportion of non-longhaulers’ comments and their sentiment, as classified by the model trained on restaurant reviews.

As a validation of these results, the same analyses were conducted for the model that was trained on laptop reviews (Figure S9 in the SM). The sentiment classification of longhauler comments showed no significant effects for parosmia (β = -0.22, SE = 0.19, *p* = .24, 95% CI = 0.56-1.15, OR = 0.80), phantosmia (β = -0.47, SE = 0.28, *p* = .08, 95% CI = 0.36-1.06, OR = 0.62), hyposmia (β = 0.16, SE = 0.20, *p* = .43, 95% CI = 0.80-1.72, OR = 1.17), or anosmia (β = 0.27, SE = 0.62, *p* = .66, 95% CI = 0.37-4.39, OR = 1.31). For the non-longhauler comments (Figure S10 in the SM), a significant effect was found for parosmia (β = -0.71, SE = 0.30, *p* = .02, 95% CI = 0.27-0.89, OR = 0.49), and hyposmia (β = -0.55, SE = 0.28, *p* = .05, 95% CI = 0.33-1.00, OR = 0.58), but not phantosmia (β = 0.01, SE = 0.51, *p* = .99, 95% CI = 0.37-2.81, OR = 1.01).

### Aim 3

To visually explore differences in food and non-food items mentioned differently in the comments of longhaulers and non-longhaulers, word-clouds were made (Figure 4). Superficially, the word-clouds appeared similar for both groups. Non-longhaulers appeared to mention “cheese”, “urine,” and “sweat” somewhat more often than longhaulers. Both groups most often mentioned “coffee,” “onion,” and “food,”, and for the non-food items, “perfume”, and “smoke.”

**Figure 4.**
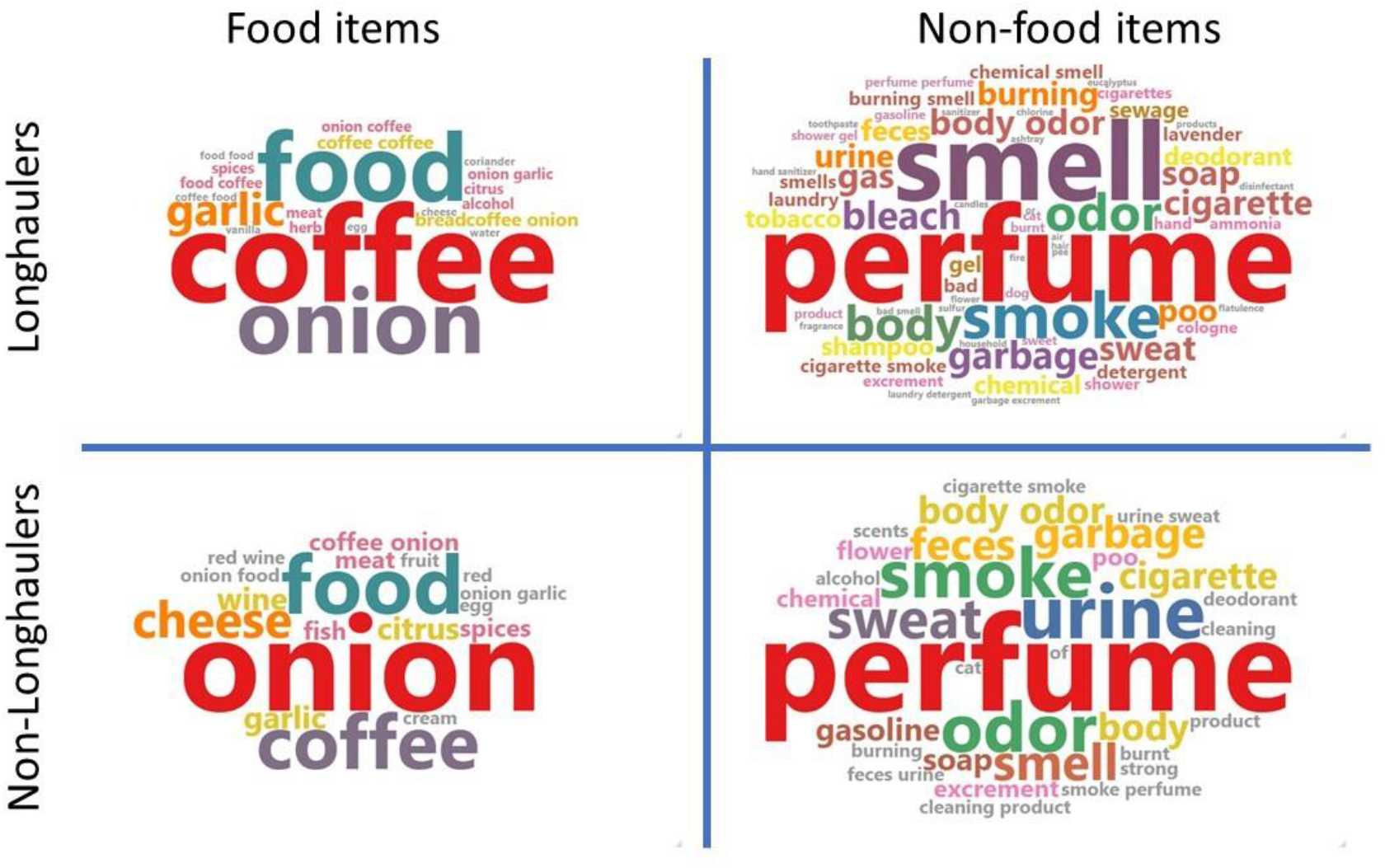
Word clouds of longhauler and non-longhauler comments Word Cloud information gathered from the participants regarding “food” and “non-food” items extracted from the comments.

Additional word clouds were created for each specific smell dysfunction (Figure 5). There does not seem to be much difference between the food-words mentioned by any of the participants. Further, most of the words seem to be consistent with the words mentioned in figure 6. However, in terms of non-food words, phantosmic participants seem to mention more words that are related to a burning sensation (e.g. “smoke”, “burning”).

**Figure 5.**
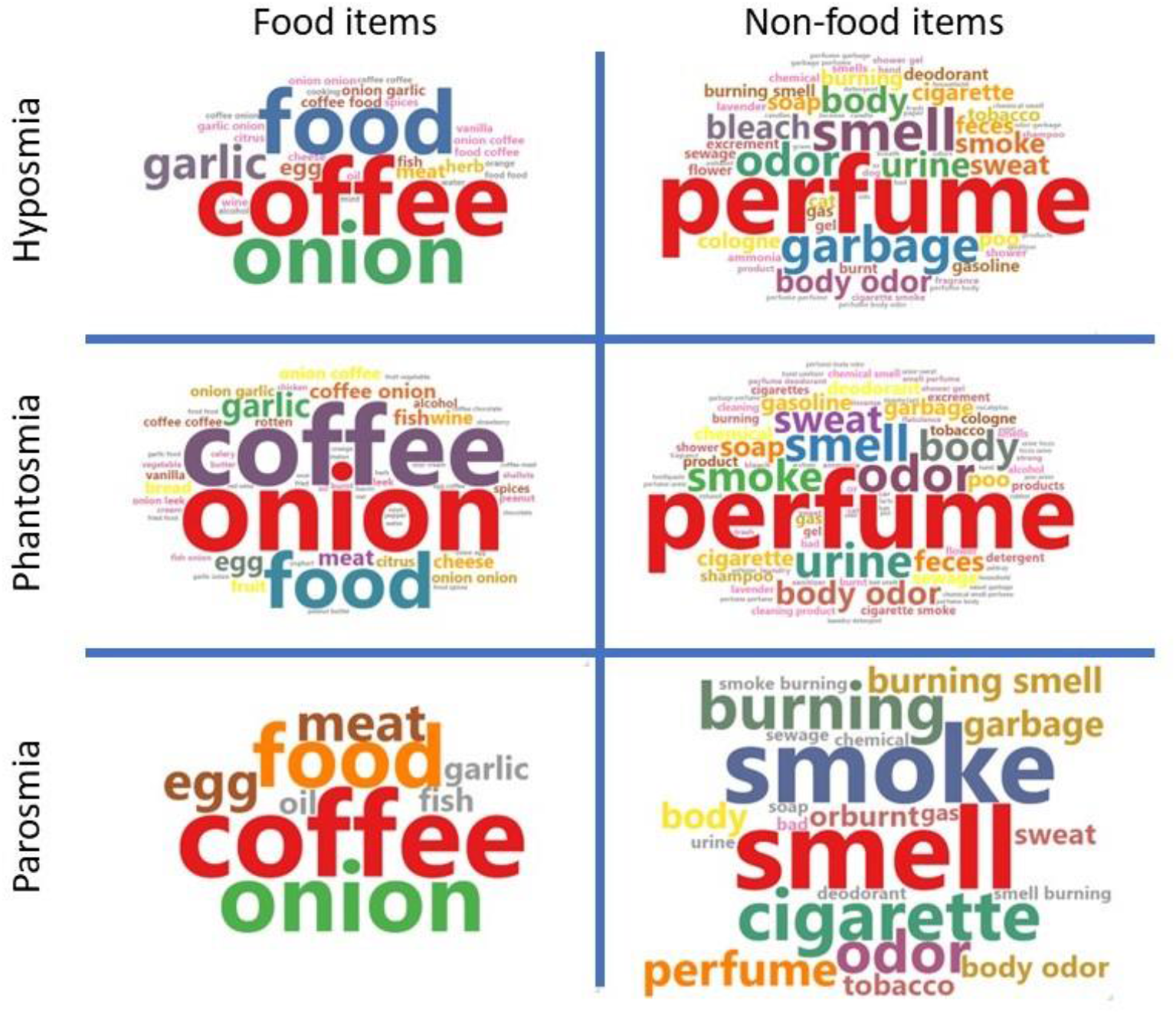
Word clouds of comments for each olfactory dysfunction Word Cloud information gathered from both longhaulers and non-longhaulers combined, regarding “food” and “non-food” items extracted from the comments grouped by specific smell dysfunction.

To follow up this visual exploration, a relative frequency analysis was conducted, comparing non-longhaulers and longhaulers. This analysis, reported in Table 3, revealed that ‘lemon’ was mentioned more often by longhaulers, whereas “wine,”‘, ‘cheese,’, ‘vinegar,’, and ‘mustard,’ were mentioned more often by non-longhaulers. For the non-food items, longhaulers more often mentioned ‘weird’ (presumably for ‘weird smells’), ‘fire,’ ‘gas,’ and ‘eucalyptus’ among other smelling objects. This is in line with the finding that longhaulers more often report parosmia (see findings Aim 1), and thus report more of these foul smelling objects in their comments, whereas non-longhaulers might report the objects that they can smell and taste again in their comments (e.g.,’wine’, ‘cheese’).

**Table 3.** List of Words That Were Reported Significantly (p < .05) More Often by Longhaulers or Non-longhaulers^b^.

|  | Log-likelihood | Occurrence |
| --- | --- | --- |
| **Food items** |  |  |
| Lemon | 7.62 | 0.78 |
| Vinegar | 9.90 | -0.50 |
| Cheese | 6.12 | -0.52 |
| Wine | 12.60 | -0.63 |
| Mustard | 4.22 | -0.76 |
| Red wine | 9.46 | -0.86 |
| **Non-food items** |  |  |
| Weird | 7.62 | 0.78 |
| Fire | 7.62 | 0.78 |
| Gas | 10.20 | 0.72 |
| Eucalyptus | 4.75 | 0.71 |
| Detergent | 6.14 | 0.64 |
| Air | 9.14 | 0.60 |
| Scent | 8.07 | 0.32 |
| Burning | 4.90 | 0.27 |
| Smoke | 4.05 | 0.24 |
| Thing | 9.40 | 0.19 |
| Very | 5.90 | 0.13 |
| Smell | 4.08 | 0.03 |

<sup>b</sup> A Larger Occurrence Value Indicates That Words Were Reported More Frequently by Longhaulers, And A Lower (Negative) Value Indicates a Higher Frequency for Non-longhaulers.

## Discussion

COVID-19 strongly impacts the life of patients, during the disease and in many cases, long afterwards. Among the more striking symptoms that impacted everyday life were those pertaining to the chemical senses, namely, taste and olfaction. The present paper focuses on the difference between participants experiencing transient and those presenting enduring chemosensory symptoms, named non-longhaulers and longhaulers, respectively. Their responses were derived from the responses to an online survey administered at two time points, during the disease and after recovery from the infection, regardless of enduring symptoms. The responses to a series of multiple-choice questions were already analysed in a previous study^1^. In this paper, we focused on the responses to open-ended questions, presented in the same surveys in which the participants could enter their own text. These allowed us to explore the emotional experiences that people went through and can be of value to understand the real impact of long-lasting olfactory symptoms in everyday life.

We considered the validity of our analyses of open-ended responses, as opposed to analyses of close-ended questions^1^. Concurrent with their results, we found that longhaulers were more likely to suffer from parosmia and hyposmia, but did not find this difference for phantosmia. We believe that this is the result of the lack of overlap between our coding and participants’ self-reports. F-scores showed that coders generally under-estimated the prevalence of smell disorders. This effect was highest in phantosmia, and may, therefore, have led to the discrepancy between ^1^ and the current study. Possible explanations for the high under-estimation of prevalence by coders are that participants did not report their whole experience in the comments, or that participants over-estimated their experiences in the multiple-choice question. This is a down-side to our approach, but overall our F-scores seem to be in a good range, and should therefore allow for a useful interpretation of the results that our natural language processing measures yielded.

Our data show that, according to our first hypothesis, longhaulers reported a significantly higher occurrence of olfactory dysfunctions, in particular, parosmia and hyposmia, than non-longhaulers did. This is in agreement with previous research that shows that olfactory dysfunctions, especially parosmia, appears to be part of long-COVID^1^. This marks the need for accurately defining the sensory experience by introducing clinical routine testing that may highlight the recovery pathway, even if partial, as well as to give feedback to patients on their clinical course. It also points towards a need for better sensory education among the population who often has no means of describing their perception accurately.

Our second aim was to link the presence of emotional distress to longhauler status by classifying comments as positive, negative or neutral. This approach takes advantage of a machine learning model, thus uncoupling classification from human evaluation. This is noteworthy, since the experience of disease is personal, and correctly classifying a patient’s experience, independently from the observer, could benefit from automated processes to assist and support clinicians. The sentiment classifier showed that comments of longhaulers were linked to a more negative tone. When clustered by symptoms, the classifier that was trained on restaurant reviews showed that the presence of parosmia or hyposmia was linked to the use of more negative words in both longhaulers and non-longhaulers. However, this was only replicated for non-longhaulers by the classifier that was trained on laptop reviews. Hence,this approach does not allow for unambiguous interpretation of sentiments in specific smell disorders. The tendency of these results are interesting nonetheless, and follow-up analysis that more specifically delves into the quality of life could shed a clearer light on the matter.

A semantic analysis of the objects mentioned within the comments suggested that the most frequently mentioned objects were similar for longhaulers and non-longhaulers concerning non-food items (e.g., “perfume”, “smoke”), while for food items, the two most frequent words were the same but switched between longhaulers and non-longhaulers. The word “onion”‘ was used most frequently by non-longhaulers and “coffee”‘ by longhaulers. Concerning differentially reported food words, “lemon”‘ was mostly reported by longhaulers and “red wine,”‘ by non-longhaulers. When looking at the specific olfactory dysfunction, the largest problem seems to arise in the non-food words mentioned by phantosmic participants. They seem to often mention words like “smoke” and “burning”.

The objects that both groups reported might be the odor objects that are most salient on one’s mind, i.e., the first objects one might think of when being prompted to come up with objects with a smell. Longhaulers may report that they no longer smell these (i.e., ‘I can no longer smell “coffee”, “perfume”, “onion”), whereas non-longhaulers may report these as the first smells that they can smell again, or the smells that they most often encounter (“I first noticed the smell of coffee again”). However, the extracted words in Aim 3 were not analyzed with regard to the direction of valence. It might be interesting to examine whether certain foods were avoided or approached during the period of smell dysfunction. Moreover, certain compounds may trigger parosmia^35^, and interestingly, the associated smells of these compounds seem to be reported quite often in the comments. Examples in this regard are the pyrazines, and their association with coffee, the disulfides with onion, the thiols with garlic or rotten eggs, and the methoxypyrazines with wine. This could present a range of verbally reliable indicators for parosmia.

The mentioning of words that describe a burning sensation in phantosmic participants is consistent with previous findings^36^. It provides confirmation of these findings in a large sample, and offers an overview of other words and sensations that may be associated with phantosmia. Although multiple hypotheses on the causes of phanstomia have been proposed, it may very well be that phanstomic sensations have varying causes^37^. The present data consists of a wide range of self-reports of these phanstomic sensations, and are therefore suitable for a follow-up analysis on specific contexts that evoke a phanstomic sensation to further the understanding of phantosmic experiences and the mechanisms that underlie them. This also highlights the value of the internet in medical research, which allows for a collection of large datasets on patients’ self-reports that may add to the understanding of olfactory dysfunctions.

The difficulty people face in discussing smells and olfactory experiences is a well-known phenomenon (e.g. ^12^). In this study, we analysed open-ended comments to gain insight into the experiences of individuals suffering from olfactory dysfunctions, that are not entirely understood or correctly named by patients suffering from these. First, we found that there is a discrepancy between the information in the open-ended comments and the close-ended multiple choice question that asks for olfactory dysfunction. This suggests that the comments contain valuable information that differs between groups and symptoms, highlighting the importance of this approach. This study emphasises the importance of considering open-ended comments to gain a more holistic understanding of participants’ experiences and perceptions. The approach presented here, i.e., manually coding short open-ended responses for different symptoms, may be used in combination with machine learning classification paradigms, to better understand patients’ concerns voiced in online settings such as in online survey research, in online patient council groups, or by using brief digital notes from general practitioners as input. This additional source of information could lead to better identification of different diagnoses, and along the line, better understanding of the different types of smell disorders.

Finally, few people mention that months after suffering from COVID-19 induced smell dysfunctioning, their smell has improved, even to a level of functioning that is better than before the onset. We have identified 13 cases of what is known as *hyperosmia*. It is currently unknown in what percentage this phenomenon occurs, and what the mechanisms behind it are. Speculatively, an increase in awareness of smells after having lost one’s sense of smell for a brief period could potentially drive attentional experience with odours - in an ‘you don’t know what you’ve got until you lose it’ way^38^. Future, large scale prevalence studies on smell disorders could investigate what percentage of people report an improved sense of smell once recovered from the smell dysfunctioning.

### Limitations of the present study

The present study has several limitations. One refers to a selection bias in this participant group: As in ^1^, we are aware that there might be a self-selection bias in completing these surveys, as participants suffering from severe symptoms may be more motivated to also complete the second survey. At the same time, participants that felt that they suffered more, may select additional answer options on the multiple choice questions and clarify their symptoms in the open comment field. Thus, this selection bias, as compared with ^1^, could go either way. This provides an opportunity to compare both ways of asking about symptoms from participants, which we included in Aim 1. Notwithstanding, we will interpret the results with caution and phrase future suggestions and implications cautiously.

## Data Availability

The pre-registration of our analyses, as well as all data and analysis scripts associated with this manuscript can be found at OSF.io (https://osf.io/xv6mn/). This analysis was done as part of the Global Consortium for Chemosensory Research (https://gcchemosensr.org/).

## Author contributions

**Conceptualization:** Ilja Croijmans, Surabhi Bhutani, Keiland W. Cooper, Paloma Rohlfs Dominguez, Michael C. Farruggia, Thomas Heinbockel, Sachiko Koyama, Nick S. Menger, Vonnie Shields, and Anna D’Errico.

**Data curation:** Ilja Croijmans, Surabhi Bhutani, Michael C. Farruggia, Nick S. Menger, Arnaud Tognetti, and Veronica Pereda-Loth.

**Formal analysis:** Ilja Croijmans, Keiland W. Cooper, Thomas Heinbockel, Nick S. Menger, Arnaud Tognetti, and Vonnie Shields.

**Funding acquisition:** Carla Mucignat.

**Methodology:** Ilja Croijmans, Surabhi Bhutani, Keiland W. Cooper, Nick S. Menger, and Arnaud Tognetti.

**Project administration:** Ilja Croijmans, Surabhi Bhutani, and Sachiko Koyama.

**Resources:** Ilja Croijmans, Surabhi Bhutani, Keiland W. Cooper, Paloma Rohlfs Dominguez, Michael C. Farruggia, Thomas Heinbockel, Nick S. Menger, Carla Mucignat, Denis Pierron, Arnaud Tognetti, Vonnie Shields, Anna D’Errico, and Veronica Pereda-Loth.

**Software:** Ilja Croijmans, Keiland W. Cooper, Nick S. Menger, and Arnaud Tognetti.

**Supervision:** Ilja Croijmans, Surabhi Bhutani, Nick S. Menger, and Arnaud Tognetti.

**Visualization:** Ilja Croijmans, Keiland W. Cooper, Michael C. Farruggia, Nick S. Menger, and Arnaud Tognetti.

**Writing - original draft:** Ilja Croijmans, Paloma Rohlfs Dominguez, Nick S. Menger, and Arnaud Tognetti.

**Writing - review & editing:** Ilja Croijmans, Surabhi Bhutani, Keiland W. Cooper, Paloma Rohlfs Dominguez, Michael C. Farruggia, Thomas Heinbockel, Sachiko Koyama, Nick S. Menger, Carla Mucignat, Denis Pierron, Arnaud Tognetti, Vonnie Shields, and Anna D’Errico.

