## Supplementary material for "Giving a voice to adults with COVID-19: An analysis of open-ended comments from smell longhaulers and non-longhaulers": Coding scheme S1.docx

**Coding scheme S1. Guidelines that were used in aim 1 to code the comments regarding their olfactory dysfunction.**

- Every ‘smell change’ comment is coded by means of dummy coding in 5 smell change categories, each in one column:
  1. Parosmia (0: no; 1: yes)
  2. Phantosmia (0: no; 1: yes)
  3. Anosmia (0: no; 1: yes)
  4. Hyposmia (0: no; 1: yes)
  5. Hyperosmia (0: no; 1: yes)
  6. Recovered (0: no; 1: partial; 2: fully)
- Comments are also coded for mentioning of specific objects:
  7. Food and beverages (0: not mentioned; 1: present in comment)
  8. Non-food items (0: not mentioned; 1: present in comment)
- Coding in the present: at time of filling in the survey (i.e., ignore reports of previous changes in smell)

1. Parosmia
   - Mentioning that *something* (a source) smells different than before, generally worse than before, ‘it is like…’. Also: ‘everything smells like […]’
2. Phantosmia
   - Smelling something when a source is not present, smelling something that is not there, ‘it smells like […] (all the time)’.
3. Anosmia
   - ‘I can’t smell anything.’ ‘No smell at all’
4. Hyposmia
   - Smell is back to some extent (relative to before). ‘I can smell to some (lesser) extent’. ‘I can smell but smells/odors are faint’, or ‘smell comes and goes’ (morning/evening). ‘I can only smell [very specific real present object smell]’
5. Hyperosmia
   - Smell ability is better than before.
6. Recovered
   - Smell is back (2: yes). Smell is back to some degree/any recovery of smell (1: partial).
7. Some categories are mutually exclusive:
   - Parosmia -> no anosmia
   - Anosmia -> no hyposmia, recovery is always 0, Anosmia -> No parosmia
   - Hyposmia -> no anosmia
8. No coding of item-specific anosmia (e.g. “I cannot perceive the smell of flowers and of sage”). These items should be commented as unsure, and can be discussed later.
