## Supplementary material for "Giving a voice to adults with COVID-19: An analysis of open-ended comments from smell longhaulers and non-longhaulers": Figure S8.pdf

**Figure S8. Odds ratios from the logistic regressions examining the association between smell longhauling and prevalence of olfactory disorders.** Longhauler status: smell long- versus non-longhauler (reference category), Gender: men versus women (reference category), Translation: yes versus no (reference category).

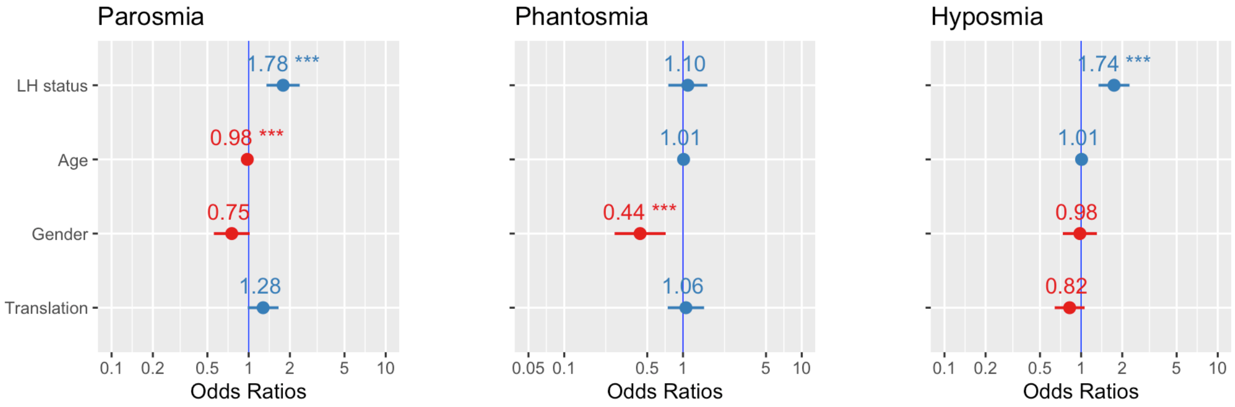
