## Supplementary material for "Giving a voice to adults with COVID-19: An analysis of open-ended comments from smell longhaulers and non-longhaulers": Figure S9.docx

**Figure S9.** **Sentiment classification from the model that was trained on laptop reviews.** Showing the proportion of comments from longhaulers that were classified as negative, neutral, and positive across all smell disorders.

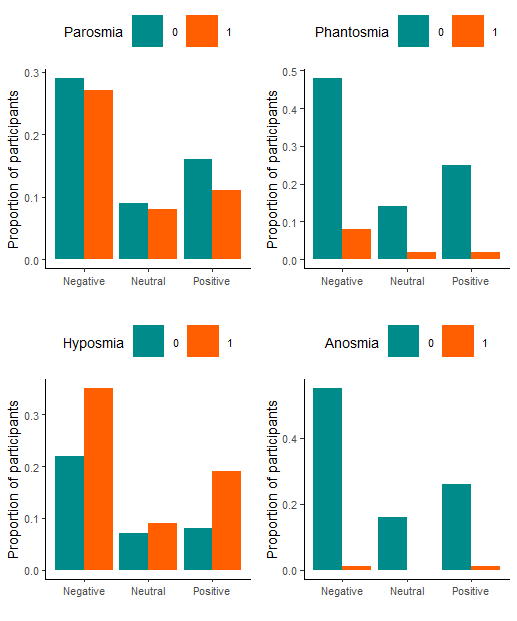
