## Supplementary material for "Giving a voice to adults with COVID-19: An analysis of open-ended comments from smell longhaulers and non-longhaulers": Figure S10.pdf

**Figure S10. Sentiment classification from the model that was trained on laptop reviews.** Showing the proportion of comments from non-longhaulers that were classified as negative, neutra, and positive across all smell disorders.

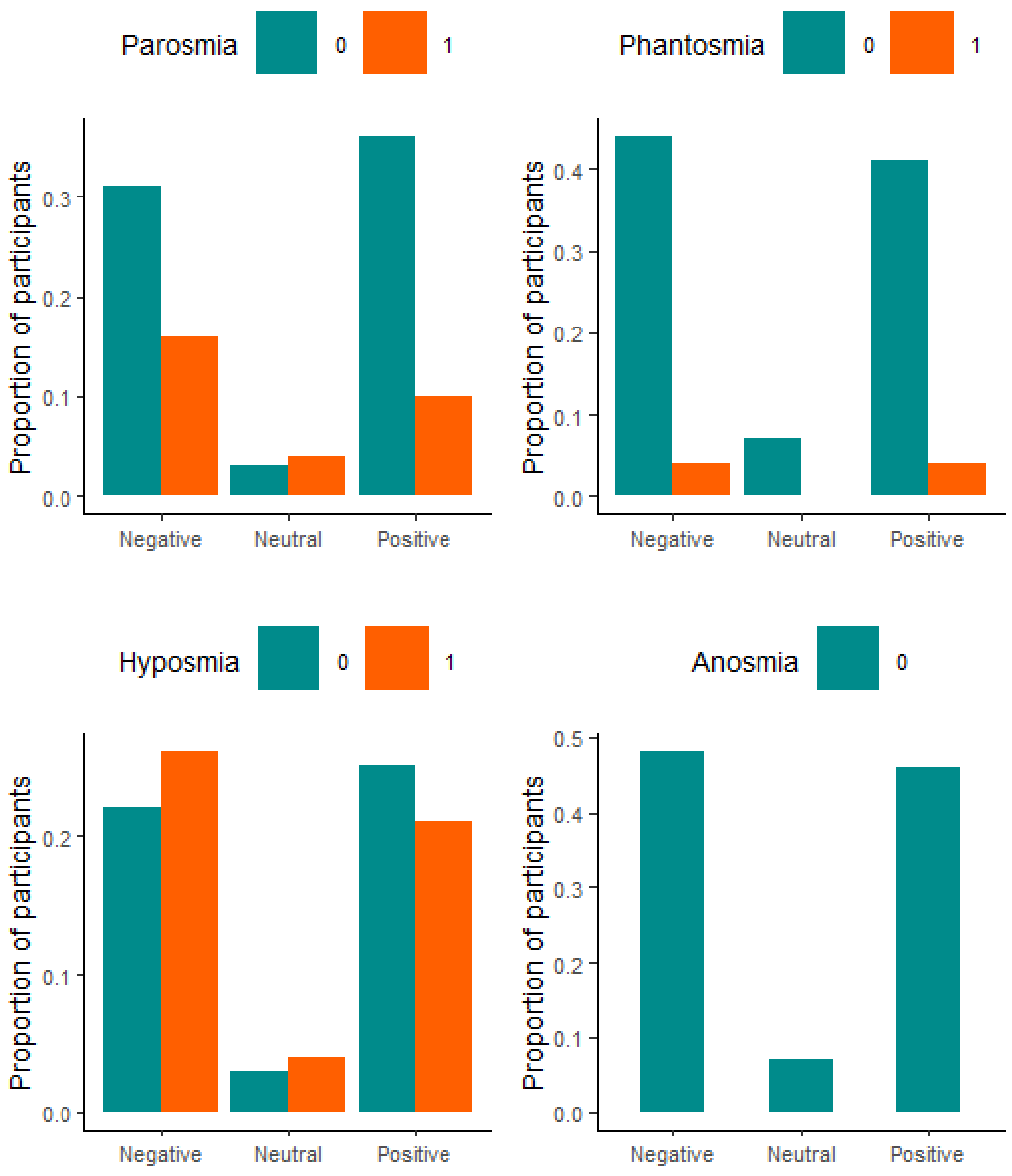
