## Supplementary material for "Giving a voice to adults with COVID-19: An analysis of open-ended comments from smell longhaulers and non-longhaulers": SM_Tables_and_Figures.docx

**Giving a voice to adults with COVID-19: An analysis of open-ended comments from smell long-haulers and non-long-haulers**

**SUPPLEMENTARY DATA**

**Table S1.**  **Confusion matrix showing the agreement between the coders (open-ended comments), and the participants’ self-report (multiple choice question) for parosmia (S1 and S2 combined)**. The cross-table shows how many comments were coded as indicating a certain smell disorder (Coder: 1) or not (Coder: 0), and the overlap with the participants’ report of having a certain smell disorder (Self-report: 1) or not (Self-report: 0). All “None”-entries were excluded from the analysis.

|  |  | Coder: 0 | Coder: 1 |
| --- | --- | --- | --- |
|  | Self-report: 0 | 1150 | 174 |
|  | Self-report: 1 | 448 | 314 |

**Table S2.**  **Confusion matrix showing the agreement between the coders (open-ended comments), and the participants’ self-report (multiple choice question) for phantosmia (S1 and S2 combined)**. The cross-table shows how many comments were coded as indicating a certain smell disorder (Coder: 1) or not (Coder: 0), and the overlap with the participants’ report of having a certain smell disorder (Self-report: 1) or not (Self-report: 0). All “None”-entries were excluded from the analysis.

|  |  | Coder: 0 | Coder: 1 |
| --- | --- | --- | --- |
|  | Self-report: 0 | 1566 | 73 |
|  | Self-report: 1 | 309 | 136 |

**Table S3.**  **Confusion matrix showing the agreement between the coders (open-ended comments), and the participants’ self-report (multiple choice question) for smell loss, i.e. anosmia and hyposmia (S1 and S2 combined).** The cross-table shows how many comments were coded as indicating a certain smell disorder (Coder: 1) or not (Coder: 0), and the overlap with the participants’ report of having a certain smell disorder (Self-report: 1) or not (Self-report: 0). All “None”-entries were excluded from the analysis.

|  |  | Coder: 0 | Coder: 1 |
| --- | --- | --- | --- |
|  | Self-report: 0 | 323 | 530 |
|  | Self-report: 1 | 250 | 987 |

**Table S4.** **Logistic regression investigating whether smell long- vs. non-longhaulers differed in terms of reported parosmia they respectively experienced.** For each variable, the estimate (β), the standard error of the mean (SE), the z statistic, and the p-value are given. The estimate of the variable *Longhauling status* is for the comparison between the longhaulers (reference category) and the non-longhaulers. The estimate of the variable *Gender* is for the comparison between men (reference category) and women. The estimate of the variable *Translation* is for the comparison between translated (reference category) or untranslated comments into English.

|  |  | ß | *SE* | z | *p* |
| --- | --- | --- | --- | --- | --- |
|  | Intercept | -0.97 | 0.15 | -6.27 |  |
|  | Longhauling status | 0.58 | 0.14 | 4.06 | <0.0001 |
|  | Age* | -0.02 | 0.005 | -4.27 | <0.0001 |
|  | Gender | -0.28 | 0.15 | -1.85 | 0.06 |
|  | Translation | 0.24 | 0.13 | 1.84 | 0.07 |

* The variable *Age* was centered.

**Table S5.** **Logistic regression investigating whether smell long- vs. non-longhaulers differed in terms of reported phantosmia they respectively experienced.** For each variable, the estimate (β), the standard error of the mean (SE), the z statistic, and the p-value are given. The estimate of the variable *Longhauling status* is for the comparison between the longhaulers (reference category) and the non-longhaulers. The estimate of the variable *Gender* is for the comparison between men (reference category) and women. The estimate of the variable *Translation* is for the comparison between translated (reference category) or untranslated comments into English.

|  |  | ß | *SE* | z | *p* |
| --- | --- | --- | --- | --- | --- |
|  | Intercept | -1.73 | 0.20 | -8.474 |  |
|  | Longhauling status | 0.09 | 0.19 | 0.49 | 0.63 |
|  | Age* | 0.01 | 0.007 | 1.51 | 0.13 |
|  | Gender | -0.83 | 0.25 | -3.29 | 0.001 |
|  | Translation | 0.06 | 0.18 | 0.31 | 0.75 |

* The variable *Age* was centered.

**Table S6.** **Logistic regression investigating whether smell long- vs. non-longhaulers differed in terms of reported hyposmia they respectively experienced.** For each variable, the estimate (β), the standard error of the mean (SE), the z statistic, and the p-value are given. The estimate of the variable *Longhauling status* is for the comparison between the longhaulers (reference category) and the non-longhaulers. The estimate of the variable *Gender* is for the comparison between men (reference category) and women. The estimate of the variable *Translation* is for the comparison between translated (reference category) or untranslated comments into English.

|  |  | ß | *SE* | z | *p* |
| --- | --- | --- | --- | --- | --- |
|  | Intercept | 0.05 | 0.14 | 0.36 |  |
|  | Longhauling status | 0.55 | 0.13 | 4.16 | <0.0001 |
|  | Age* | 0.008 | 0.005 | 1.48 | 0.14 |
|  | Gender | -0.02 | 0.15 | -0.14 | 0.89 |
|  | Translation | -0.19 | 0.13 | -1.51 | 0.13 |

* The variable *Age* was centered.

**Figure S7.** **Odd-ratios from the logistic regressions examining the association between smell longhauling and prevalence of olfactory disorders.** Longhauler status: smell long- versus non-longhauler (reference category), Gender: men versus women (reference category), Translation: yes versus no (reference category).

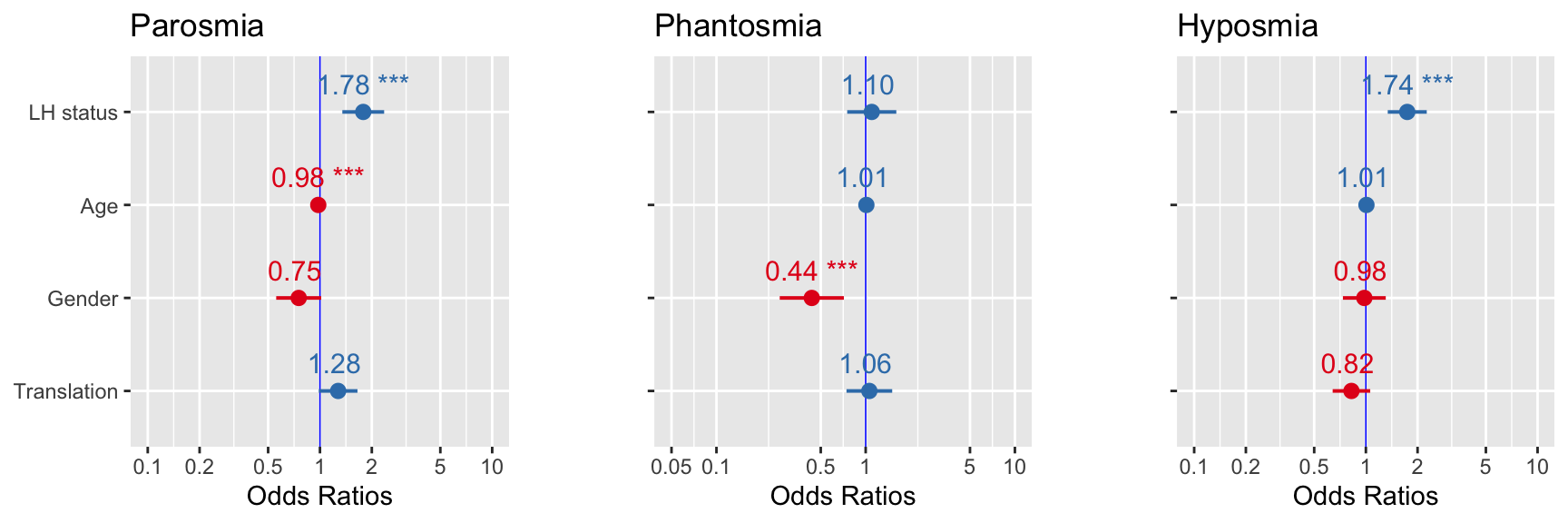

**Figure S8.** **Sentiment classification from the model that was trained on laptop reviews.** Showing the proportion of comments from longhaulers that were classified as negative, neutral, and positive across all smell disorders.

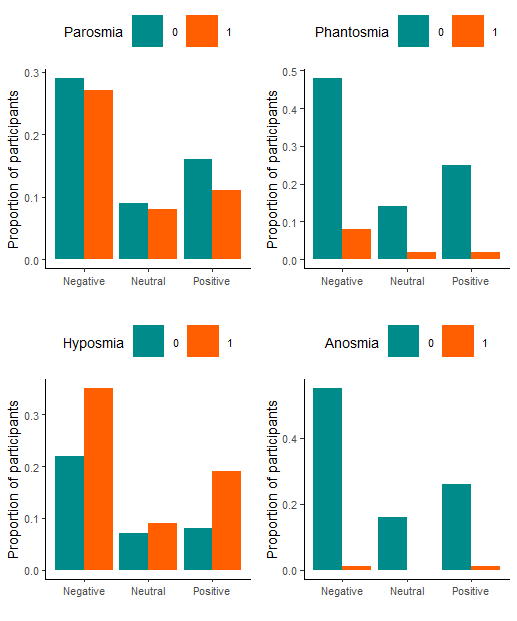

**Figure S9.** **Sentiment classification from the model that was trained on laptop reviews.** Showing the proportion of comments from non-longhaulers that were classified as negative, neutra, and positive across all smell disorders.

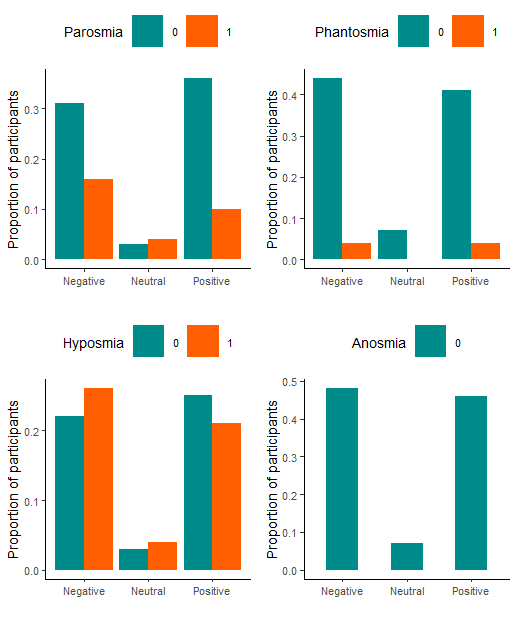
